# No compliment for complement: Kidney biopsies of patients with malignant nephrosclerosis show no complement activation

**DOI:** 10.1101/2023.01.27.23284929

**Authors:** Elisa M. Alba Schmidt, Nicola Tomas, Tobias Huber, Thorsten Wiech, Tilman Schmidt, Ulrich Otto Wenzel

## Abstract

The complement system represents an ancient part of innate immunity and is comprised of multiple serum proteins. In several pathological conditions, including atypical hemolytic syndrome (aHUS), complement activation is contributing to disease progression. Therefore, pharmacological targeting of complement is a viable therapeutic strategy in this population.

Recent data suggest complement activation in patients suffering from malignant hypertension and stenosing arteriosclerosis also called malignant nephrosclerosis (MNS). Since MNS and aHUS also share histopathological similarities therapeutic complement inhibition became a discussion in MNS.

To eliminate the pitfall of confusing complement trapping with activation in the kidney we used the proximity ligation assay to detect, visualize and quantify assembled complement convertases of the different pathways in renal tissue diagnosed with MNS or aHUS. We compared them to a control group of patients presenting with thin basement membrane syndrome (TBMD). Activation of the alternative and the classical pathways were examined by glomerular and vascular quantification of C3b/Bb and C4b/C2b, respectively.

We found an overactivation of the alternative pathway only in patients suffering from aHUS, while complement activation was not altered in MNS when compared with TBMD. Thus our results cannot contribute to the hypothesis that complement inhibition may be an efficient therapeutic strategy for patients presenting end-organ damage due to stenosing arteriosclerosis. The use of complement inhibition in these patients cannot be recommended.

## INTRODUCTION

Patients with severe hypertension and signs of end-organ damage, e.g., acute renal injury and mild signs of thrombotic microangiopathy, are labeled to suffer from “malignant hypertension” or, more correctly, stenosing arteriosclerosis. Some patients might have mild clinical and histological signs of thrombotic microangiopathy in the renal vasculature, which has been linked to mechanical stress on the endothelium. Although the prognosis of malignant hypertension has improved with modern anti-hypertensive medication, worsening renal function is still observed. Improving these undesirable outcomes depends on a better understanding of the pathogenesis. Increasing evidence indicates that hypertension and hypertensive end-organ damage are not only mediated by hemodynamic injury but that inflammation plays a vital role and contributes to this disease^1,2^. The complement system is an ancient part of innate immunity comprising multiple serum proteins and cellular receptors protecting the host from a hostile microbial environment and maintaining tissue and cell integrity. It also plays a central role in regulating adaptive immunity. Thus, complement activation may drive the pathology of hypertension and hypertensive injury through its impact on innate and adaptive immune responses aside from direct effects on the vasculature^3,4^. Indeed, recent experimental data support a role for complement in hypertensive end-organ damage, suggesting that complement inhibition should be used in patients with malignant nephrosclerosis^5–7^.

Today the analysis of the complement system’s contribution to renal diseases is primarily based on the immunohistochemical detection of deposited complement activation products in tissue and the detection of consumption of complement components. To eliminate the pitfall of complement trapping in injured tissue with activation in the kidney, we used the proximity ligation assay (PLA) to detect, visualize and quantify assembled complement convertases of the different pathways in renal tissue diagnosed with malignant hypertension^8^.

## RESULTS

The complement system is activated via three pathways: alternative, classical, and lectin. Spontaneous, continuous low-level default hydrolysis of C3 to C3b and the formation of the alternative C3 convertase by binding and cleavage of factor B (C3bBb) characterize the alternative complement pathway. The classical complement pathway is activated when C1q binds to antibodies and undergoes a conformational change to an activated protease. Cleavage of C4 and C2 forms the C3 convertase of the classical pathway. The lectin and classical pathways share the same C3 convertase comprising C2b and C4b (C4bC2b). We used the proximity of C3b and Bb to identify the alternative, and C2b and C4b to identify the classical/lectin pathway, as shown in figure 1.

**Figure 1.**
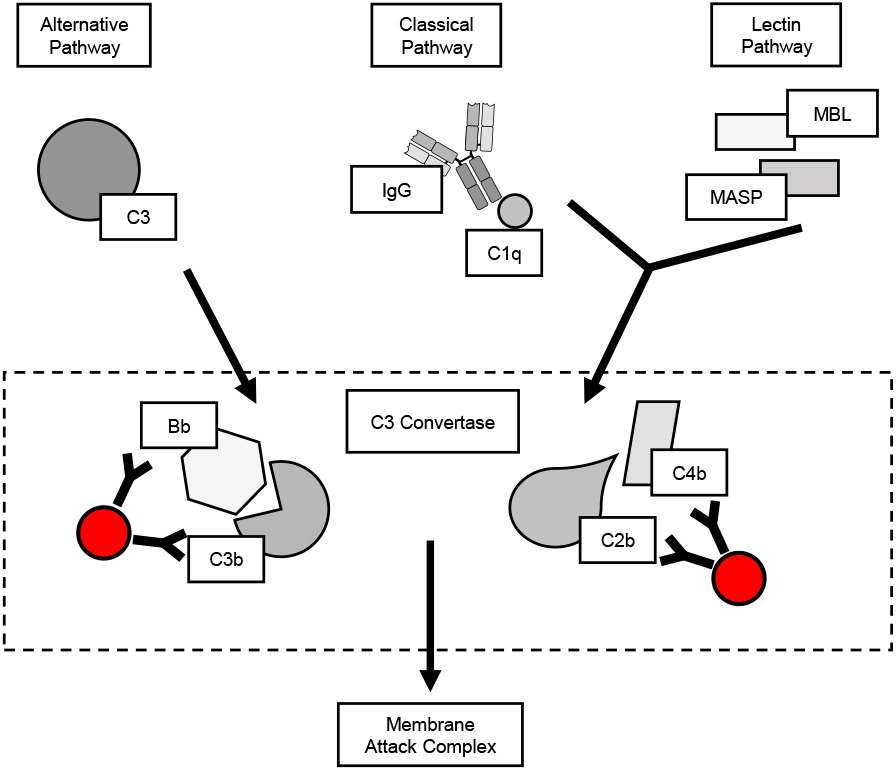
Activation of the complement system by the alternative or the classical / the lectin pathway pathway can be distinguished by the different C3 convertases. While the alternative C3 convertase is formed by C3b and Bb, the classical/lectin C3 convertase comprises C2 and C4b.

We applied the PLA in biopsies of patients with malignant nephrosclerosis (n=11, serum creatinine 5.3±1.2 mg/dl, age 42±2.9). Malignant nephrosclerosis was diagnosed in hypertensive patients with decreased renal function and narrowing of interlobular arterial branches and afferent arterioles with intima fibrosis and onion-like intimal scarring. As controls, we used biopsies diagnosed with thin basement membrane syndrome and normal renal function (n=9, serum creatinine 0.8±0.07, age 47±6). To detect complement activation as a positive control, we used patients with histopathological signs of thrombotic microangiopathy, which were positively tested for genetic alterations in more than 70% of the cases. This cohort was declared to suffer from atypical hemolytic uremic syndrome (n=11, serum creatinine 4,0±1.3, age 40±11).

Representative glomerular images of PLAs of kidney biopsies from patients with malignant nephrosclerosis, thin basement membrane syndrome, and aHUS with proximity resulting in black dots are shown in figure 2. After quantification, no significant differences were found for the alternative (C3b/Bb), nor for the classical/lectin (C2bC4b) pathway when comparing glomerular signals from controls and patients with malignant nephrosclerosis, as shown in figure 2a. This finding contrasts with significantly higher glomerular signals for the alternative complement activation (C3bBb) in the aHUS cohort. Since intrarenal vessels are the disease-defining lesion in malignant nephrosclerosis, we also counted the vascular PLA reaction. However, similar to the glomerular activity, no significant difference was found between the the control group and malignant nephrosclerosis while only alternative activation of complement was significantly higher in aHUS patients, as shown in figure 2b. These data show that none of the three complement pathways is activated in kidney biopsies from patients with malignant nephrosclerosis.

**Figure 2.**
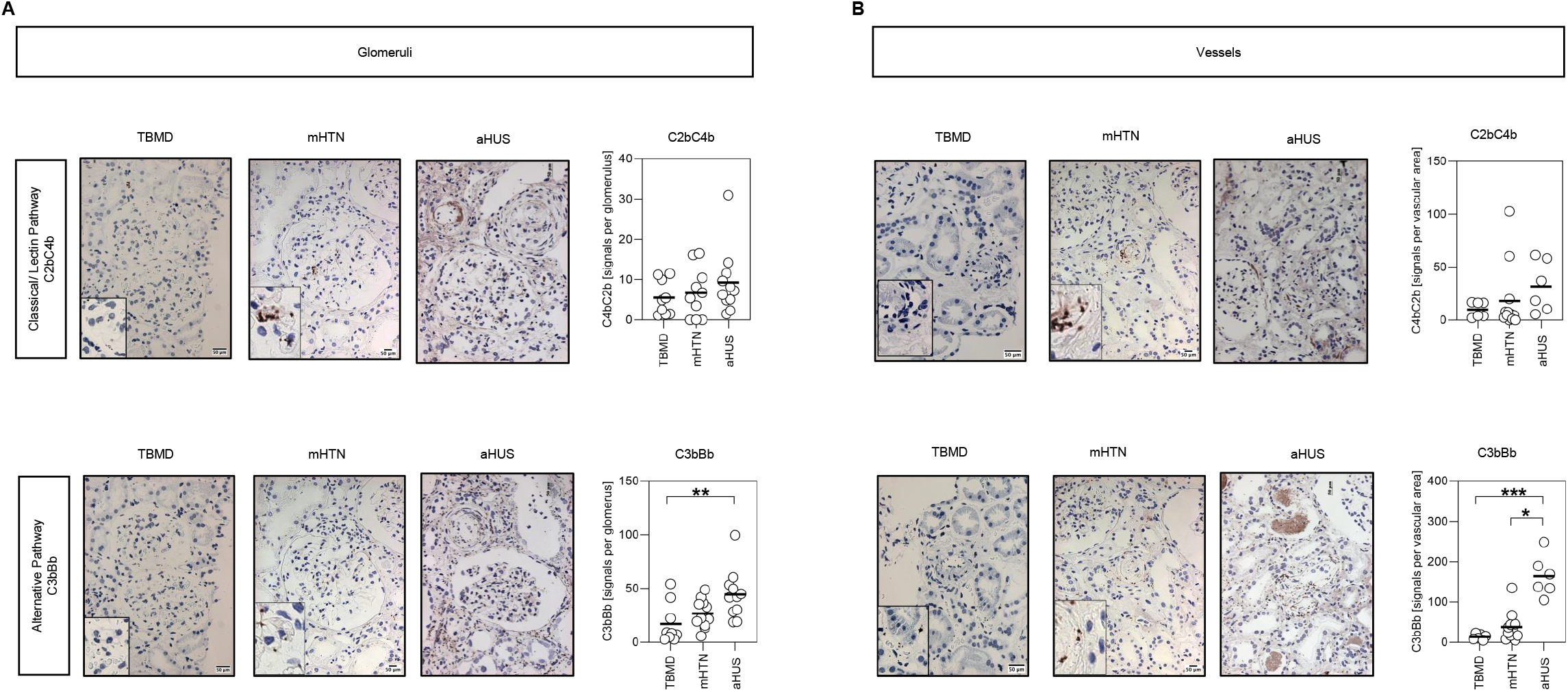
A shows representative glomeruli of patients with TBM (n=9), mHTN (n=11), and aHUS (n=11) stained for the alternative (C3bBb) C3 convertase and the classical/lectin C3 convertase (C2C4b). B shows representative vessels of the same cohorts with TBMD and mHTN stained for the alternative (C3bBb) C3 convertase and the classical/lectin convertase (C2C4b). TBMD: Thin Basement Membrane Disease; mHTN: malignant Hypertension; aHUS: atypical hemolytic uremic syndrome.

## DISCUSSION

The histologic changes found in malignant nephrosclerosis are more correctly called stenosing arteriosclerosis, defined by the narrowing of interlobular arterial branches and afferent arterioles with intima fibrosis and onion-like appearance of intimal scarring^4^. Patients presenting with malignant nephrosclerosis are often young and have a short history of uncontrolled hypertension. The mechanisms causing the transition from high blood pressure to malignant nephrosclerosis are unknown^4^. The shared clinical and histopathological features between malignant nephrosclerosis and atypical hemolytic syndrome strongly suggest a role for complement also in the development of malignant nephrosclerosis and point to complement inhibition as a potential therapeutic strategy in malignant hypertension. Uncontrolled complement activation in the alternative pathway is strongly linked with the pathogenesis of aHUS.

In 2017 Timmermans and colleagues published a study indicating complement activation in patients with malignant nephrosclerosis. Their data suggest that complement defects may be the culprit of disease, especially in patients progressing to end-stage renal disease despite blood pressure control^5^. In a second publication, Timmermans demonstrated that hypertension-associated thrombotic microangiopathy induced C5b-9 deposition on endothelial cells in an ex-vivo approach^6^. In the same direction, Zhang et al. studied complement components in serum, urine, and renal tissue of patients with malignant nephrosclerosis. They found aberrant complement alternative pathway dysregulation in these patients. The changes in the alternative pathway were associated with the activity, severity, and renal outcomes of malignant nephrosclerosis^7^. However, the data are descriptive and only suggest but do not prove activation of the alternative pathway in patients with malignant nephrosclerosis. They cannot differentiate between trapped or deposited complement fragments and complement activation, which causes renal injury. These studies used kidney biopsies to diagnose malignant nephrosclerosis or hypertension-associated thrombotic microangiopathy and most of those had signs of thrombotic microangiopathy.

Interestingly in all three studies, the authors found genetic risk mutations for aHUS^5–7^. In 2018 Timmermans found recurrent TMA in seven of 11 donor kidneys after transplantation^6^. Genetic alterations, signs of TMA, and recurrence after transplantation might favor atypical hemolytic uremic syndrome as the correct diagnosis in these patients. In addition, negative genetic testing cannot rule out the participation of the complement system in this disease. We diagnosed malignant nephrosclerosis in patients with typical signs of stenosing arteriosclerosis that lack genetic risk alerations. In contrast, aHUS was diagnosed only with histopathological signs of thrombotic microangiopathy. Most patients were positively tested for genetic risk mutations.

In the present study, we applied the PLA in renal biopsies to answer whether the complement system is activated in patients with malignant nephrosclerosis. In summary, using the proximity ligation assay, our data demonstrate that none of the three complement pathways is activated in patients with malignant nephrosclerosis. We strongly suggest that the current evidence does not allow complement inhibition in patients with malignant hypertension and malignant nephrosclerosis. Our data is supported by a study by Larsen, who genetically analyzed 100 hypertensive patients whose kidney biopsies were restricted to classic hypertension-associated lesions without glomerular microthrombi. In this study, no genetic alterations were found^9^. In addition, our study demonstrates activation of the alternative complement pathway in kidneys suffering from aHUS. For that, in the future, PLA could be a helpful tool in renal pathology to rule out aHUS and the need for eculizumab in patients with severe hypertension, renal failure, thrombotic microangiopathy, and a histologic finding compatible with malignant nephrosclerosis or aHUS with severe hypertension.

## DISCLOSURE

TS received fees from Alexion and Novartis. This support did not influence the research work or the content of this manuscript.

## Data Availability

All data produced in the present study are available upon reasonable request to the authors

## REFERENCES

1. Drummond GR, Vinh A, Guzik TJ, Sobey CG. Immune mechanisms of hypertension. Nat Rev Immunol. 2019;19(8):517–532. doi:10.1038/s41577-019-0160-5

2. Hengel FE, Benitah JP, Wenzel UO. Mosaic theory revised: inflammation and salt play central roles in arterial hypertension. Cell Mol Immunol. 2022;19(5):561–576. doi:10.1038/s41423-022-00851-8

3. Ruan CC, Gao PJ. Role of Complement-Related Inflammation and Vascular Dysfunction in Hypertension. Hypertension. 2019;73(5):965–971. doi:10.1161/HYPERTENSIONAHA.118.11210

4. Wenzel UO, Kemper C, Bode M. The role of complement in arterial hypertension and hypertensive end organ damage. Br J Pharmacol. 2021;178(14):2849–2862. doi:10.1111/bph.15171

5. Timmermans Sameg, Abdul-Hamid MA, Vanderlocht J, et al. Patients with hypertension-associated thrombotic microangiopathy may present with complement abnormalities. Kidney International. 2017;91(6):1420–1425. doi:10.1016/j.kint.2016.12.009

6. Timmermans Sameg, Abdul-Hamid MA, Potjewijd J, et al. C5b9 Formation on Endothelial Cells Reflects Complement Defects among Patients with Renal Thrombotic Microangiopathy and Severe Hypertension. JASN. 2018;29(8):2234–2243. doi:10.1681/ASN.2018020184

7. Zhang Y, Yang C, Zhou X, et al. Association between thrombotic microangiopathy and activated alternative complement pathway in malignant nephrosclerosis. Nephrology Dialysis Transplantation. 2021;36(7):1222–1233. doi:10.1093/ndt/gfaa280

8. Person F, Petschull T, Wulf S, et al. In situ Visualization of C3/C5 Convertases to Differentiate Complement Activation. Kidney International Reports. 2020;5(6):927–930. doi:10.1016/j.ekir.2020.03.009

9. Larsen CP, Wilson JD, Best-Rocha A, Beggs ML, Hennigar RA. Genetic testing of complement and coagulation pathways in patients with severe hypertension and renal microangiopathy. Mod Pathol. 2018;31(3):488–494. doi:10.1038/modpathol.2017.154

