## Supplement for "No compliment for complement: Kidney biopsies of patients with malignant nephrosclerosis show no complement activation"

**Material and Methods**

PATIENT SELECTION

From the archive of the institute of pathology, section for nephropathology of the University Hospital Hamburg-Eppendorf, 9 biopsies of patients histopathological diagnosed with thin basement membrane disease, 11 with malignant hypertension and 10 with aHUS were selected. Thin Basement Membrane Disease (TBMD) was histopathologically diagnosed by a experienced nephro-pathologist. Malignant hypertension (mHTN) was diagnosed in kidney biopsies of hypertensive patients with decreased renal function in which narrowing of interlobular arterial branches and afferent arterioles with intima fibrosis and onion-like intimal scarring was the dominant finding. Patients with atypical hemolytic uremic syndrome (aHUS) were identified by renal biopsies showing typical signs of thrombotic microangiopathy, lacking histological signs of severe hypertensive damage. Eight of 11 patients were positively tested for genetic risk mutations.

Patient characteristics

|  | TBMD | mHTN | aHUS |
| --- | --- | --- | --- |
| Age | 47±6 | 42±3 | 40±11 |
| Gender (male/female) | 3/6 | 7/4 | 5/6 |
| Serum creatinine (mg/dl) | 0.8±0.07 | 5.3±1.2 | 4,0±1.3 |
| Genetic risk mutation | n.d. | n.d. | 8/11 (72,7%) |

Genetic risk mutation of aHUS patients:

| Mutation | n |
| --- | --- |
| CFH | 4 |
| CFI | 2 |
| CFHR3/1 deletion | 1 |
| CFHR3/CFH hybrid | 1 |
| CFHR3/CFHR4 hybrid | 1 |

The study was approved by the local ethics committees at Hamburg (Ethics commission Hamburg, WF 011/15). All work was carried out in accordance with the Declaration of Helsinki.

PROXIMITY LIGATION ASSAY

Paraffin sections (1-2 µm) were deparaffinized, rehydrated and pretreated with proteinase (Sigma-Aldrich) for 30 min. at 40°C. Endogenous peroxidases were quenched with H2O2 (Duolink® In Situ Detection Reagents Brightfield) and samples were blocked using normal horse serum (Vector Laboratories) for 10 min. at 37°C. Antibodies against the classical C3/C5 convertase components C2 (mouse, 1:10; Santa Cruz sc373809) and C4b (rabbit, 1:500; Abcam ab181241) as well as against the alternative C3/C5 convertase C3b (mouse, 1:1000; Abcam ab11871) and CFB (rabbit, 1:20; Proteintech 10170-1-AP) were each subsequently applied overnight at 4°C. Antibodies against IgG (mouse, 1:7500; Jackson ImmunoResearch Laboratories 209-005-088) and C1q (rabbit, 1:1500; Dako A0136) were applied for 30 min. at 37°C. Secondary anti-mouse (Duolink® In Situ PLA® Probe Anti-Mouse MINUS) and anti-rabbit (Duolink® In Situ PLA® Probe Anti-Rabbit PLUS) antibodies were applied according to protocol, followed by the detection brightfield kit (Duolink® In Situ Detection Reagents Brightfield) for visualization. Nuclear staining was performed using Mayer’s Hematoxylin.

Manual Analysis was performed with a ZEISS AXIO Scope.A1 microscope, biopsies were analyzed with the Plan-APOCHROMAT 40x/0,95 corr. Ocular. Signal counting was performed manually assisted via the application for iOS “WBC Counter” (by Kazuyoshi Sasaoka version 1.1.0). The glomerular area was measured with the ZEN Vision program for Microsoft, pictures were taken via the ZEISS AxioCam MRc (TV 2/3 “C 0.63x).

A mean of 7.0, 5.3, and 4.1 glomeruli and 4, 2.3 and 1.4 vessels per kidney biopsy were evaluated for TBMD, mHTN and aHUS, respectively.

STATISTICAL ANALYSIS

For the statistical analysis, GraphPad Prism 9.2.0 was used. Data were presented as mean and individual data points. For analysis of significance, we used Kruskal-Wallis test for multiple comparisons.
